# Diastolic Age: A Cardiac Biological Clock Derived from Echocardiography and the PREVENT Heart Failure Risk Score

**DOI:** 10.64898/2026.04.15.26350995

**Authors:** Gracia Fahed, Nicholas Cauwenberghs, Everton Santana, Rongrong Chen, Bettia Celestin, Bruna Gomes, Sarah A. Short, Megan K. Carroll, Tatsuya Miyoshi, Kevin M. Alexander, Svati H. Shah, Shai S. Shen-Orr, Attila Kovács, Melissa M. Daubert, Tatiana Kuznetsova, Karima Addetia, Federico Asch, Kenneth W. Mahaffey, Pamela S. Douglas, Francois Haddad

## Abstract

**Background:** Among cardiac measures, diastolic parameters demonstrate the earliest and most consistent age-related changes. This can be leveraged to develop a continuous left ventricular (LV) Diastolic Age from routine echocardiographic parameters. Analogous to how epigenetic clocks weight molecular markers against mortality risk, we calibrated Diastolic Age by weighting echocardiographic features against the validated PREVENT–Heart Failure (HF) risk score.

**Methods:** We analyzed 1,952 participants from the Project Baseline Health Study (median age 50 [36–64] years, 54% female). The measure was derived using partial least-squares regression anchored on PREVENT-HF and calibrated within a healthy reference subgroup. External validation was performed in the WASE (n=1,708) and Stanford Cardiovascular Aging (n=313) cohorts. Associations with ASE-defined LV diastolic dysfunction (LVDD), epigenetic clocks, and major adverse cardiovascular events (MACE) were examined.

**Results:** Diastolic Age correlated strongly with chronological age (r=0.78) with robust external validation (WASE r=0.76; Stanford r=0.82; calibration slopes ≈1.0). It increased progressively across grades of diastolic dysfunction and discriminated LVDD with an AUC of 0.89 (95% CI 0.87–0.92), and was independently associated with hypertension, diabetes, and elevated C-reactive protein. While correlated with the Levine (r=0.76) and Horvath (r=0.41) epigenetic clocks, residual analyses indicated that Diastolic Age captures a distinct cardiac-specific dimension of biological aging. Over median follow-up of 4.2 years, it independently predicted MACE (HR 2.30, 95% CI 1.70–3.18), with accelerated diastolic aging across all age groups among those with events. Discrimination was comparable to ASE-defined LVDD (C-index 0.83 vs. 0.82).

**Conclusions:** Diastolic Age provides a continuous, echocardiography-derived measure of cardiac biological aging that complements categorical diastolic grading and epigenetic aging clocks, and independently predicts cardiovascular outcomes.

## Introduction

Chronological age alone does not fully capture biological aging, which varies widely across individuals.^1^ Biological clocks, which can be based on epigenetic, proteomic, or physiological markers, aim to quantify this variation.^2–9^ Epigenetic clocks were among the first to be developed, with Horvath calibrating DNA methylation patterns against chronological age across multiple tissue types.^2^ Levine later expanded this framework by weighting methylation markers against a mortality-derived phenotypic age from the National Health and Nutrition Examination Survey (NHANES) cohort, linking biological aging more directly to health outcomes.^3^ These clocks hold significant promise in improving disease risk prediction, guiding preventive interventions and evaluating treatment responses beyond what chronological age alone provides. However, they require specialized blood draws and costly laboratory analysis. More recently, attention has shifted to organ-specific clocks, particularly in cardiovascular medicine, that are easily calculated using routine clinical data. For example, McClelland et al. developed a coronary artery calcium (CAC)-based vascular age calibrated on incident cardiovascular events.^10^ Cardiac magnetic resonance, echocardiography and electrocardiography derived clocks have also been described, primarily aligned to chronological age.^11–13^

Among cardiovascular measures, diastolic function parameters demonstrate the earliest and most consistent age-related changes.^14^ Data from the Framingham Heart Study, the FLEMENGHO study, and others confirm that early diastolic mitral annular velocity and transmitral filling patterns are the most sensitive cardiac markers of aging.^14,15^ Diastolic dysfunction often precedes symptomatic heart failure and independently predicts adverse outcomes,^15^ yet its clinical assessment remains categorical.^14^ A continuous diastolic age could complement current classification systems by capturing risk gradients that categorical grading of left ventricular diastolic dysfunction (LVDD) may miss.

A key challenge in constructing any biological clock is selecting the calibration target (**Table 1**). Chronological age is universally available but does not capture risk factors or early-stage cardiovascular diseas^2^; clinical outcomes offer robust anchors but are limited by low event rates, especially in younger individuals^10^. Risk score based calibration, as used by the Levine clock^3^ and cardiovascular risk scores^16,17^, offers a middle path. The American Heart Association (AHA) Predicting Risk of Cardiovascular Events (PREVENT)–Heart Failure (HF) risk score, which integrates age, sex, traditional risk factors, and their interactions, provides a suitable standard for a cardiac-specific clock.^18^

**Table 1.**
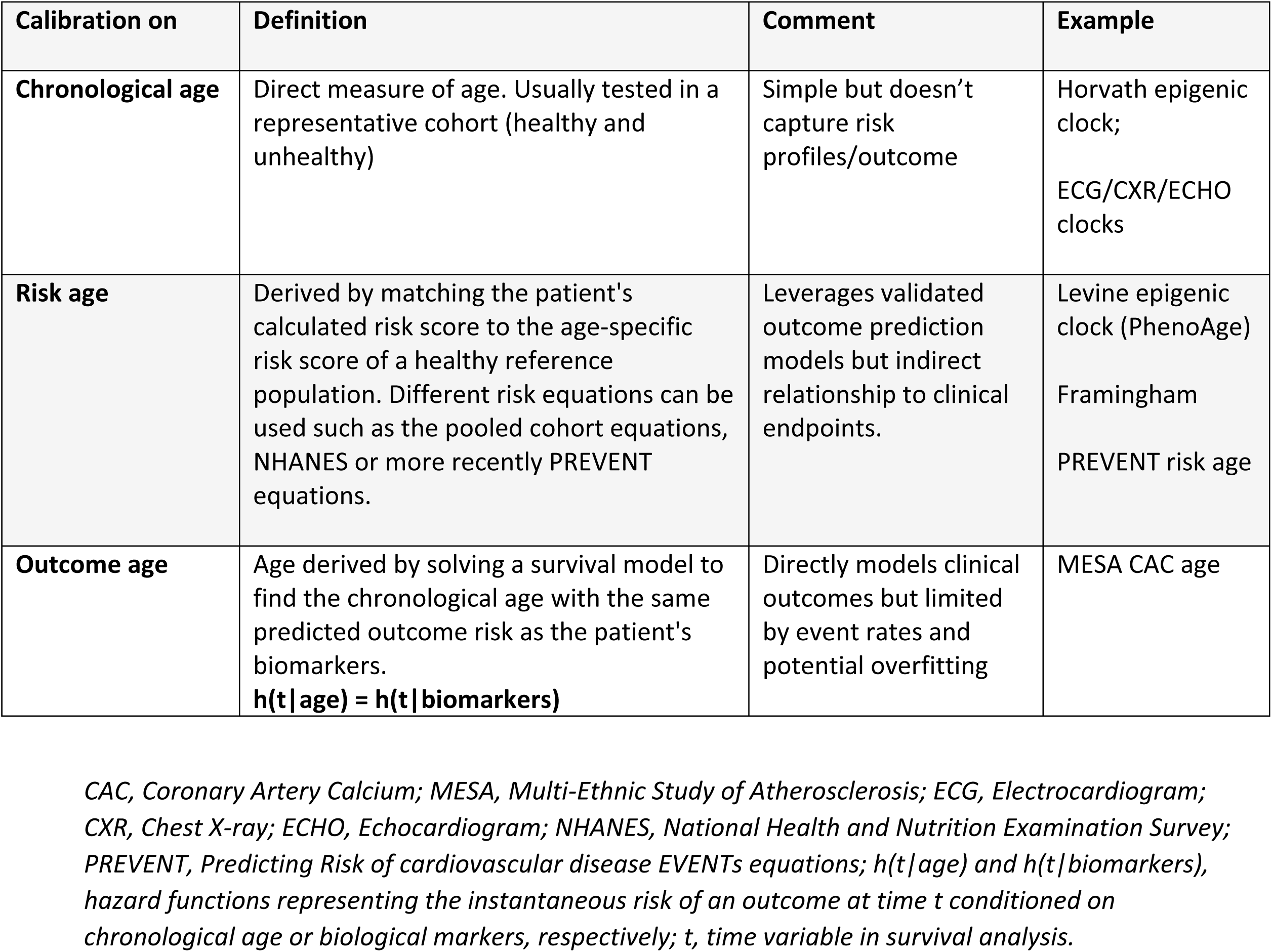
Calibration Methods for Biological Age.

In this study, we derived a left ventricular (LV) Diastolic Age from routine echocardiographic parameters, weighted by the PREVENT-HF risk score and calibrated within a healthy reference subgroup. We assessed its relationship with the American Society of Echocardiography (ASE)-defined LVDD criteria and with established epigenetic clocks, and we explored whether it independently predicts cardiovascular outcomes. Data was leveraged from the Project Baseline Health Study (PBHS)^19^, with external validation in the World Alliance of Societies in Echocardiography (WASE) reference cohort^20^ and a Stanford Cardiovascular Aging cohort.

## Methods

### Study Population

The design of the PBHS has been previously described.^19^ The PBHS enrolled 2,502 participants across multiple US sites (Stanford University, Duke University, and the California Health and Longevity Institute). The study was approved by the Stanford University and Duke University Institutional Review Boards, and participants provided written informed consent. For this sub-study, we included patients who underwent a baseline echocardiography and had diastolic measurements available. We excluded those in whom diastolic dysfunction could not be assessed reliably (moderate or severe mitral valve disease, atrial fibrillation or flutter at the time of echocardiography, or a paced rhythm). The resulting overall study cohort included 1,952 participants **(Figure 1)**.

**Figure 1.**
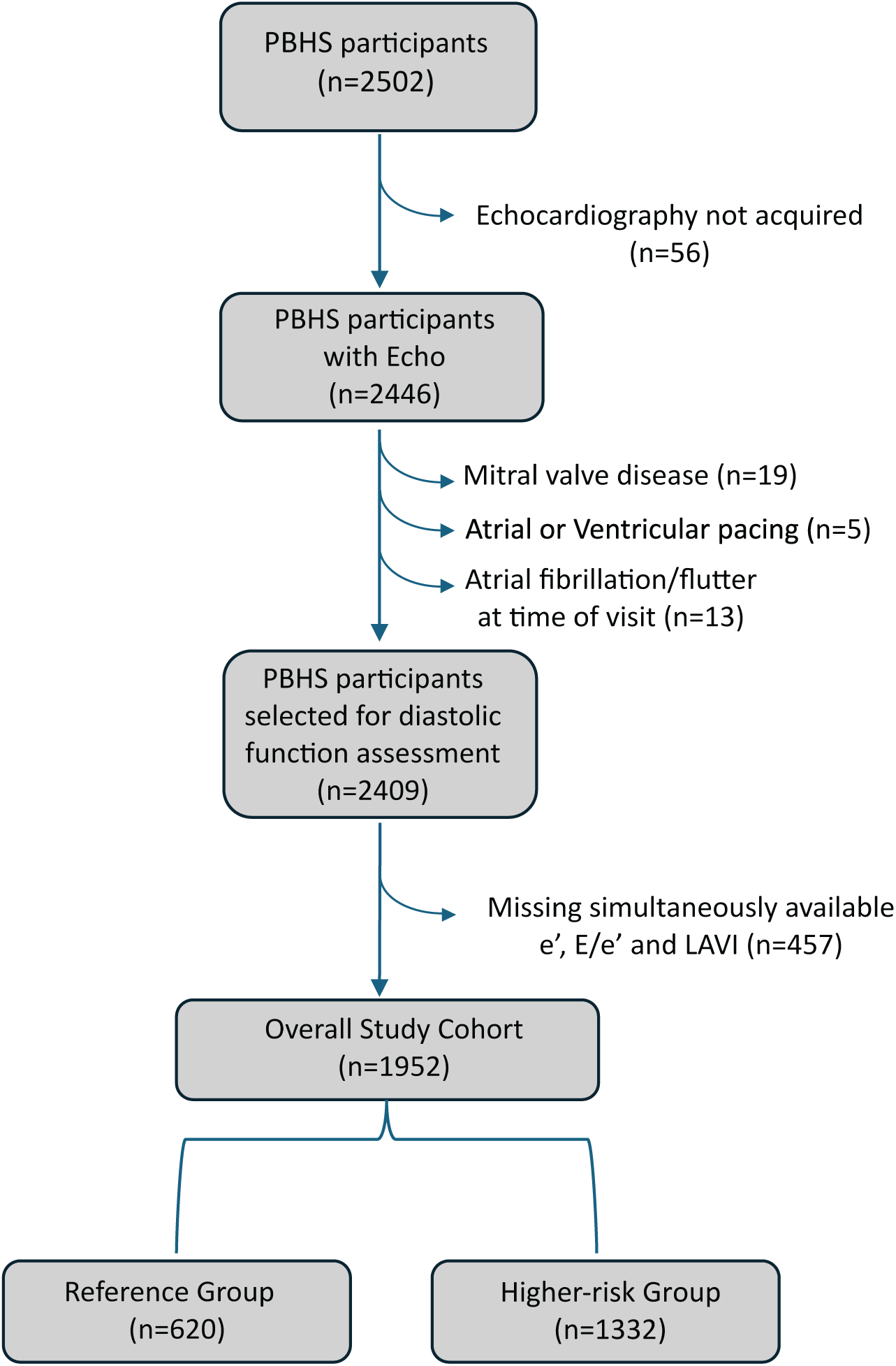
Flow Chart of the Project Baseline Health Sub-Study Population. For the reference group, participants with established diseased (cardiovascular, pulmonary, kidney, autoimmune disease or active cancers), major cardiovascular risk factors and metabolic syndrome were excluded. *Abbreviations: LAVI, left atrial volume index; PBHS, project baseline health study*.

Within this cohort, we defined a healthy reference group (n= 620) for model calibration, consisting of individuals with no current or prior history of cardiovascular, pulmonary, kidney, or autoimmune diseases or active cancers; and no major atherosclerotic risk factors including hypertension, diabetes mellitus, obesity (BMI ≥ 30 kg/m^2^ **(Figure 1)**. The PREVENT-HF risk score was calculated at baseline in participants without established cardiovascular disease who had complete data for all required variables with values within the acceptable ranges specified by the original PREVENT equations (n=1,299).^18^

### Echocardiography

Echocardiographic measurements included M-mode, 2-dimensional, pulsed-wave Doppler and tissue Doppler imaging. Parameters included LV linear dimensions, LV mass, septal and lateral e’, early and late diastolic peak velocities of mitral inflow ratio (E/A), average E/e’, left atrial volumes indexed to body surface area (LAVI), LA reservoir strain (LARS), peak tricuspid regurgitation velocity (pTRV) and estimated right ventricular systolic pressure (RVSP).^21^ Chamber quantification and assessment of diastolic function was performed by Duke Imaging Core Laboratory, blinded to patients’ clinical data, using the Digisonics software platform. A total of 3 to 5 signals were measured and averaged. Reproducibility was excellent with intraclass correlation coefficients of 0.94 for LV end-diastolic volume, 0.99 for LV mass, 0.96 for LV ejection fraction, and 0.94 for LAV.

Left ventricular diastolic dysfunction (LVDD) was classified and graded according to the 2025 ASE guidelines.^14^ Among the supporting criteria, only LAVI and LARS were available. Therefore, when both primary grading criteria—tissue Doppler e′, E/e′ ratio, or pTRV/RVSP—were discordant or inconclusive, we defined an “indeterminate grade” category to enable more granular analysis. This category, while not formally included in the ASE recommendations, was introduced to preserve analytic resolution and better characterize transitional diastolic patterns within the cohort.

### Epigenetic clocks

In the PBHS cohort, two epigenetic clocks (Horvath and Levine) were measured to assess biological aging.^4^ The Horvath pan-tissue clock utilizes DNA methylation patterns across 353 CpG sites regressed on chronological age.^2^ The Levine clock (PhenoAge) predicts phenotypic age based on methylation at 513 CpG sites derived from a combination of chronological age and blood biomarkers (albumin, creatinine, glucose, C-reactive protein, lymphocyte percent, mean cell volume, red cell distribution width, alkaline phosphatase, and white blood cell count) that were selected based on their ability to predict mortality risk in the NHANES cohort.^3^ DNA methylation profiling was performed using the Illumina Infinium MethylationEPIC 850K BeadChip array from genomic DNA extracted from blood samples. The detailed laboratory protocols, bioinformatics pipeline, and quality control procedures for methylation analysis have been previously described.^4^

### Outcome Assessment

Major adverse cardiovascular events (MACE) were reported by participants or confirmed by study coordinators using pre-specified visit case report forms, collected centrally, and independently adjudicated by 2 blinded investigators. The primary end point was a composite of cardiovascular death, atherosclerosis-related (myocardial infarction, stroke, peripheral arterial disease), incident heart failure, new-onset atrial fibrillation and cardiovascular intervention (coronary revascularization, new pacemaker implantation, valve replacement).

### Validation cohorts

External validation cohorts included two independent cohorts: the WASE reference cohort and the Stanford Cardiovascular Aging cohort. The WASE cohort has been well described previously^20^; and for this study, we included individuals without obesity or hypertension who had complete diastolic parameters available (n=1708; median age of 45 years; 49% female). The Stanford Cardiovascular Aging cohort included individuals without cardiovascular disease or risk factors (n=313; median age 56 years; 56% female).

### Statistical Analysis

Continuous variables were summarized as median with interquartile range (IQR), and categorical variables as frequencies and percentages. The overall statistical pipeline is summarized in **Figure 2**.

**Figure 2.**
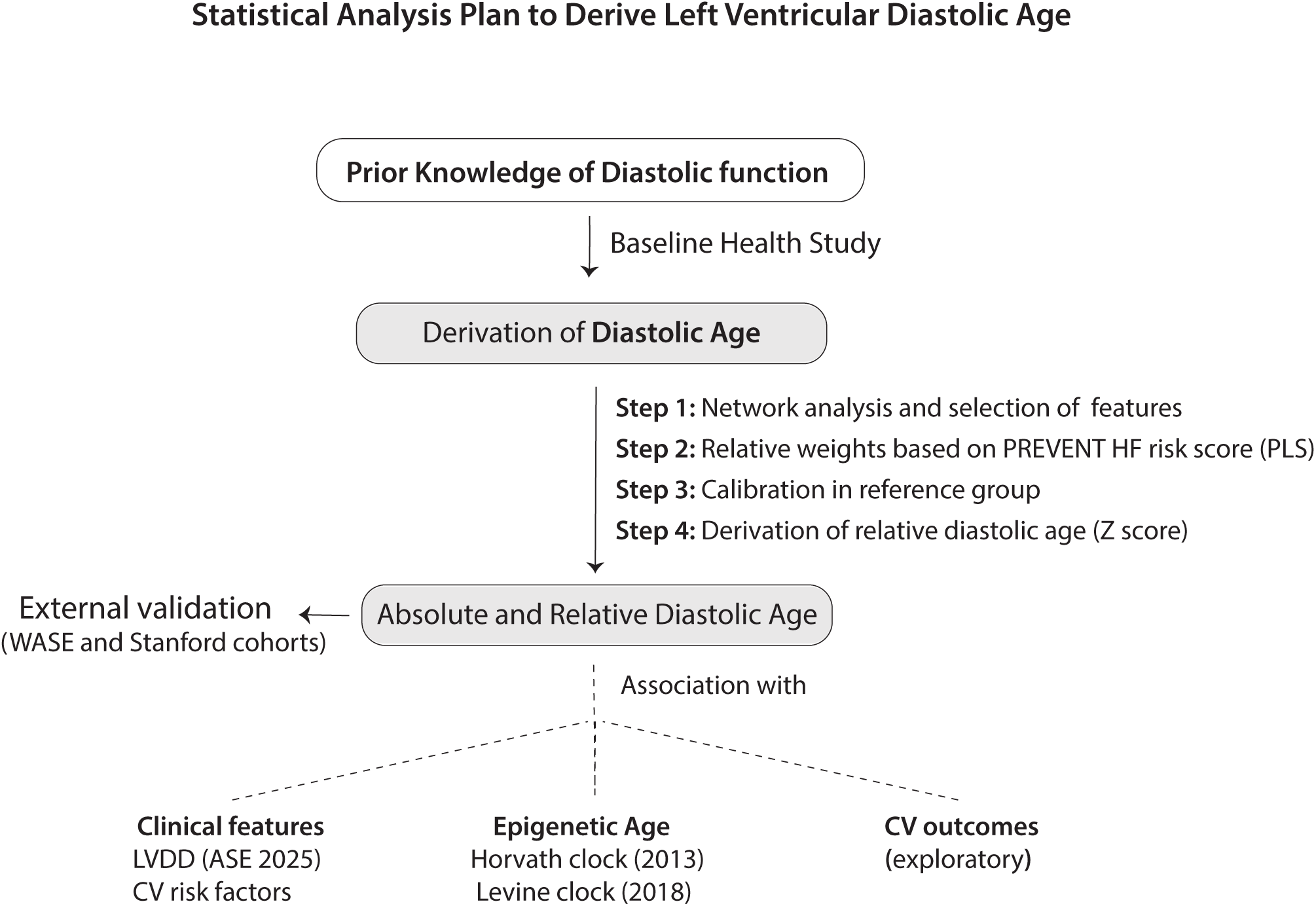
Statistical Analysis Plan of The Study. First part was dedicated for derivation and validation of diastolic age and a second part for analysis of the associations with risk factors, ASE classification of LVDD, epigenetic clocks and cardiovascular outcomes. *Abbreviations: ASE, American Society of Echocardiography; CV, cardiovascular; HF, heart failure; LVDD, left ventricular diastolic dysfunction; PLS, partial least squares*.

#### Network Analysis

To characterize the relationships among diastolic indices, chronological age, and the PREVENT-HF risk score, we performed a directed network analysis incorporating restricted cubic spline (RCS) terms with three knots to capture non-linear relationships.^22^ For each variable pair, the RCS framework selected the optimal functional form (linear or non-linear) while adjusting for multiple comparisons using the Holm correction. Model performance was evaluated using Nagelkerke’s R²,^23^ with thresholds of R² >0.08 for linear and >0.01 for logistic models. The network was visualized using a spring-layout algorithm to reflect relational strength. Hierarchical clustering using Ward’s linkage and Euclidean distance was applied to confirm grouping patterns, with dendrogram height (0–1) representing normalized dissimilarity between clusters.^24^

#### Diastolic Age Derivation

Relative weights for echocardiographic parameters were derived using partial least squares (PLS) regression, anchored on the PREVENT-HF risk score (logit) as the target variable (**Figure 2**).^25^ PLS was chosen over standard regression given the inherent correlation among diastolic predictors (e.g., e′ and E/e′). This approach prioritized diastolic features according to their association with heart failure risk rather than chronological age alone. The model included four predictors: lateral and septal e′ velocities, average E/e′ ratio, and LAVI. The first PLS latent factor explained 97% of the captured variance, with subsequent factors contributing minimally (<3%); a single-factor model was therefore retained.

The Diastolic Age equation was then calibrated within a healthy reference subgroup by aligning predicted values with chronological age using weighted least-squares regression, with chronological age and sex as candidate predictors. We derived both absolute Diastolic Age (in years) and relative Diastolic Age (as a Z-score) (**Figure 2**). The model was validated in the WASE and Stanford cohorts by assessing Pearson correlation and calibration slope. Sub-analyses were conducted by sex in both cohorts and by race in WASE cohort. For comparison and completion, we also tested alternative models: one substituting LARS for LAVI, and a second using Doppler indices alone.

#### Diastolic Age Analysis

Diastolic Age was first compared between the reference group and the remainder of the cohort, followed by stratification by major cardiovascular risk factors. Logistic regression assessed the independent contribution of individual risk factors to elevated Diastolic Age and its corresponding Z-score. We then evaluated how Diastolic Age discriminated LVDD according to ASE 2025 criteria^14^ and examined age-adjusted distributions across LVDD grades. Associations between Diastolic Age and epigenetic aging clocks (Horvath and Levine) were analyzed using correlation and residual analyses.

For outcome assessment, an exploratory analysis was performed beginning with Kaplan–Meier survival curves across Diastolic Age quantiles, followed by restricted survival tree analysis (depth of 3) to identify data-driven risk thresholds, and multivariable Cox proportional hazards models to estimate adjusted hazard ratios for predicting MACE. We compared two Cox models: ASE 2025-defined LVDD versus Diastolic Age >95th percentile (or Z-score >1.96) as predictors, with discrimination assessed via bootstrapping. Both models were adjusted for the same covariates. All statistical analyses were conducted using Python version 3.11.5, with statistical significance defined as p < 0.05.

## Results

### Study Population Characteristics

The clinical and echocardiographic characteristics of the overall cohort and reference group are presented in **Table 2**. Median age of the 1,952 participants was 50 [36-64] years; 56% females; 36% with hypertension; and 15% with type II diabetes mellitus. Compared to the overall cohort, the reference group was younger (39 [29-54] years, p<0.001) and free of CVD risk factors, yet maintained similar racial and ethnic representation.

**Table 2:** General Characteristics of The Study Population.

|  | **Overall study cohort**<br>**(n = 1952)**<br>(Median[IQR] or n (%)) | **Healthy reference**<br>**group (n = 620)**<br>(Median[IQR] or n (%)) |
| --- | --- | --- |
| ***Demographics*** |  |  |
| Age, y | 50 [36-64] | 39 [29-54] |
| Female, n (%) | 1098 (56) | 366 (59) |
| Body mass index, kg/m<sup>2</sup> | 27.2 [23.9-32.1] | 24.3 [22.2-26.3] |
| Systolic BP, mm Hg | 121 [111-133] | 113 [106-122] |
| Diastolic BP, mm Hg | 75 [69-82] | 71 [66-77] |
| ***Race*** |  |  |
| White, n (%) | 1221 (63) | 389 (63) |
| Black or African American, n (%) | 337 (17) | 57 (9) |
| Asian, n (%) | 197 (10) | 100 (16) |
| American Indian or Alaska native | 24 (1) | 6 (1) |
| Native Hawaiian or pacific<br>islander | 20 (1) | 7 (1) |
| Other, n (%) | 153 (8) | 61 (10) |
| ***Medical history*** |  |  |
| Current smoker, n (%) | 290 (15) | - |
| Hypertension, n (%) | 694 (36) | - |
| Diabetes mellitus type 2, n (%) | 294 (15) | - |
| History of CAD, n (%) | 75 (4) | - |
| COPD, n (%) | 63 (3) | - |
| Auto-immune disease, n (%) | 93 (5) | - |
| ***Biochemistry*** |  |  |
| HbA1c, % | 5.5 [5.2-5.8] | 5.3 [5.1-5.5] |
| Total cholesterol, mg/dL | 183 [157-209] | 182 [159-203] |
| LDL cholesterol, mg/dL | 97 [76-121] | 96 [78-114] |
| HDL cholesterol, mg/dL | 55 [45-69] | 62 [49-74] |
| Triglycerides, mg/dL | 107 [76-162] | 87 [64-124] |
| Creatinine serum, mg/dL | 0.84 [0.72-0.97] | 0.82 [0.71-0.93] |
| eGFR, mL/min/1.73 m<sup>2</sup> | 95 [82-107] | 101 [90-113] |
| CRP, mg/L | 1.26 [0.61-3.10] | 0.68 [0.40-1.3] |
| Uric acid, mg/dL | 5.0 [4.1-5.9] | 4.6 [3.8-5.5] |
| ***Conventional echocardiography*** |  |  |
| LV mass index, g/m<sup>2</sup> | 68 [59-79] | 64 [56-74] |
| LV ejection fraction, % | 59 [56-60] | 59 [56-60] |
| ***LV diastolic function*** |  |  |
| E/A ratio | 1.2 [0.9-1.5] | 1.4 [1.1-1.8] |
| e' septal, cm/s | 8.4 [6.8-10.6] | 10.5 [8.4-12.1] |
| e' lateral, cm/s | 11.3 [8.8-14.7] | 14.2 [11.3-16.6] |
| E/e' ratio | 7.5 [6.2-9.3] | 6.4 [5.5-7.6] |
| LA volume index, ml/m<sup>2</sup> | 26.8 [22.6-31.8] | 26.5 [22.8-31.1] |

Data are presented as median [interquartile range 25<sup>th</sup>- 75<sup>th</sup>] or n (%). *BP*, blood pressure; *CAD*, coronary artery disease; *COPD*, chronic obstructive pulmonary disease; *HbA1c*, glycated hemoglobin; *LDL*, low-density lipoprotein; *HDL*, high-density lipoprotein; *eGFR*, estimated glomerular filtration rate; *CRP*, C-reactive protein; *LV*, left ventricular; *E/A ratio*, ratio of early (E) to late (A) ventricular filling velocities; *e'*, early diastolic mitral annular velocity; *E/e' ratio*, ratio of early mitral inflow velocity to early diastolic mitral annular velocity; *LA*, left atrial.

### Left Ventricular Network

Diastolic parameters, chronological age, and the PREVENT-HF risk score were strongly interrelated illustrated by the network analysis (**Figure 3**). The PREVENT-HF logit showed the strongest relationships with chronological age (R² = 0.81), followed by lateral e′ (0.51), septal e′ (0.49), E/A ratio (0.33), E/e′ ratio (0.31), and peak TR velocity (0.15). Associations with LARS (R² = 0.05) and LAVI (0.03) were substantially weaker. Among diastolic variables, the strongest associations were observed among Doppler-derived parameters. Most associations were non-linear, as indicated by significant p-values for non-linearity after Holm correction. A complete summary of pairwise relationships is provided in **Supplemental Table 1**. Hierarchical clustering confirmed the grouping of related variables.

**Figure 3.**
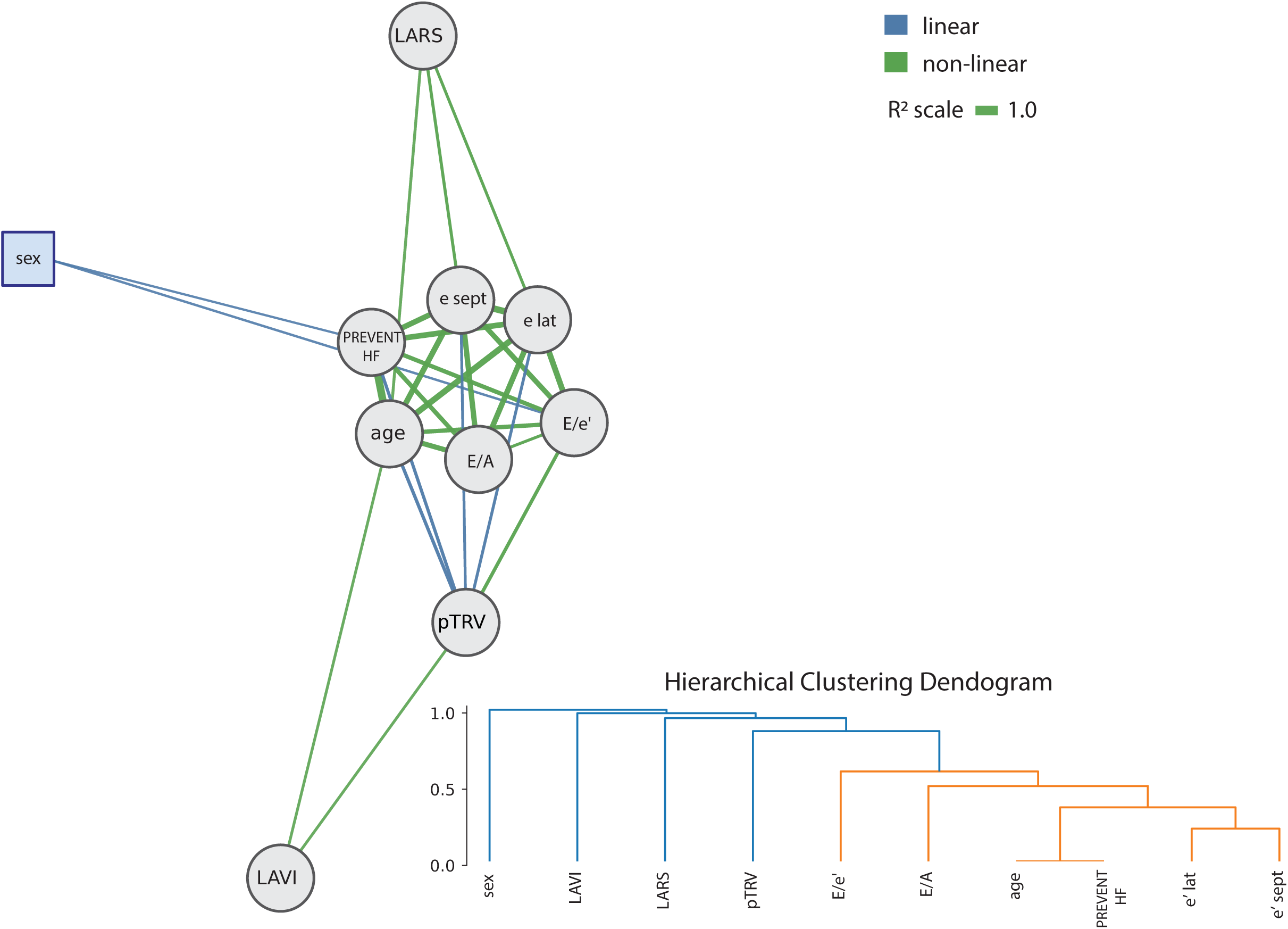
Network Analysis and Hierarchical Clustering between Age, PREVENT HF risk score and Diastolic Parameters. In the network, thicker lines represent stronger association between two variables. In the dendrogram, lower branching heights (i.e. closer to 0) indicate greater similarity between variables. *Abbreviations: E/A, ratio of early to late diastolic filling velocities; E/e’, ratio of mitral inflow E velocity to mitral annular e’ velocity; e lat, lateral early diastolic mitral annular velocity; e sept, septal early diastolic mitral annular velocity; LARS, left atrial reservoir strain; LAVI, left atrial volume index; pTRV, peak tricuspid regurgitation velocity*.

### Left Ventricular Diastolic Age Derivation and Validation

PLS analysis determined the relative weights and variable importance in projection (VIP) of diastolic parameters (**Supplemental Table 2**). The first latent factor achieved a model R² = 0.56 and captured 97% of the total explained variance. Among predictors, lateral and septal e′ velocities had the highest contribution (VIP > 1.0), followed by E/e′ ratio (VIP = 0.91) and LAVI (VIP = 0.46). For comparison, an elastic-net model yielded similar descriptive performance but substantially down-weighted E/e′, inconsistent with established physiological and outcome-based evidence.^14^ Given inherent multicollinearity among diastolic predictors, PLS was selected to extract orthogonal components that yield more stable, physiologically plausible coefficients (**Supplemental Table 3**). After calibration in the healthy reference group, the PBHS Diastolic Age equation was:

**Diastolic Age = 88.73 – 2.44 (lateral e′) – 3.47 (septal e′) + 2.49 (E/e′ average) + 0.25 (LAVI).**

Diastolic Age correlated strongly with chronological age in the reference group (r=0.78, p<0.001; **Figure 4A**), with stable variance across the predicted range. The corresponding Z-score distribution is displayed in **Figure 4B**.

**Figure 4.**
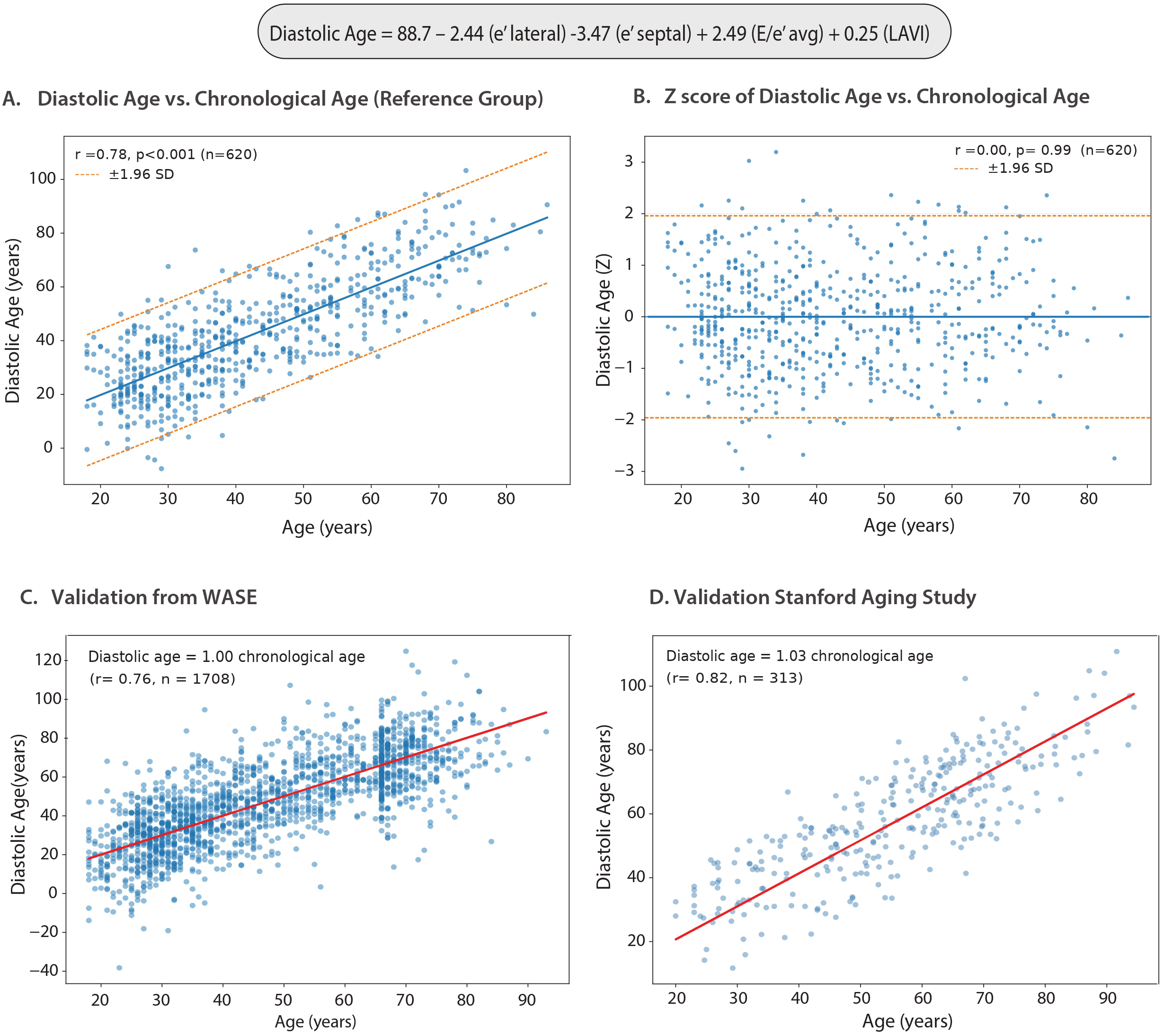
Derivation and Validation of Diastolic Age. Distribution of Diastolic Age as an absolute measure in years (Panel A) and as a Z-score-based measure (Panel B) in the Project Baseline Health Study reference group. Validation of Diastolic Age in the WASE cohort (Panel C) and the Stanford Cardiovascular Aging cohort (Panel D).

In two independent validation cohorts, Diastolic Age demonstrated robust generalizability. In the WASE cohort (**Supplemental Table 4**), Diastolic Age correlated strongly with chronological age (r=0.76, p<0.001) with excellent calibration (slope=1.00). The Stanford cohort (**Supplemental Table 5**) showed similarly strong performance (r=0.82, p<0.001). Sex-and race-specific analyses (**Supplemental Figures 1–3**) revealed consistent associations with only minor calibration differences across racial groups.

Alternative Diastolic Age models using LARS or Doppler-only measures are presented in **Table 3**. All models had R² >0.95 between them. The LARS model was not selected as the primary model due to a higher percentage of missing data.

**Table 3.** Diastolic Age equations derived from the PBHS.

| **Models** | **Diastolic Age Equations** | **R<sup>2</sup>** | **95<sup>th</sup> percentile in the reference group** |
| --- | --- | --- | --- |
| Doppler with LAVI | $88.7 - 2.44 e' \text{ (lateral)} - 3.47 e' \text{ (septal)} + 2.49 E/e' \text{ avg.} + 0.25 \text{ LAVI}$ | 0.56 | 78.3 years |
| Doppler with LARS | $102.0 - 2.28 e' \text{ (lateral)} - 3.28 e' \text{ (septal)} + 2.57 E/e' \text{ avg.} - 0.30 \text{ LARS}$ | 0.56 | 77.4 years |
| Doppler only | $96.9 + 2.50 e' \text{ (lateral)} - 3.56 e' \text{ (septal)} + 2.56 E/e'$ | 0.55 | 77.5 years |

*e' (lateral), lateral mitral annular early diastolic velocity; e' (septal), septal mitral annular early diastolic velocity; E/e' avg., average ratio of early mitral inflow velocity to mitral annular early diastolic velocity; LAVI, left atrial volume index; LARS, left atrial reservoir strain.*

### Clinical Associations with Diastolic Age

Cumulative frequency plots showed distinct Diastolic Age distributions between the reference group and the remainder of the cohort, with minimal overlap indicating clear separation of cardiac aging profiles (**Figure 5A**). Risk-stratified plots showed progressive rightward shifts across risk categories: subjects with neither diabetes nor hypertension had the lowest diastolic ages, followed by diabetes only, hypertension only, and the highest values in those with both (**Figure 5B**). Associations between risk factors and both absolute and relative Diastolic Age are shown in **Figure 5C**. Predictors of higher Diastolic Age included female sex, higher age, history of cardiovascular disease, hypertension, higher systolic blood pressure, lower HDL cholesterol, higher HbA1c, higher C-reactive protein, and Black or African American race.

**Figure 5.**
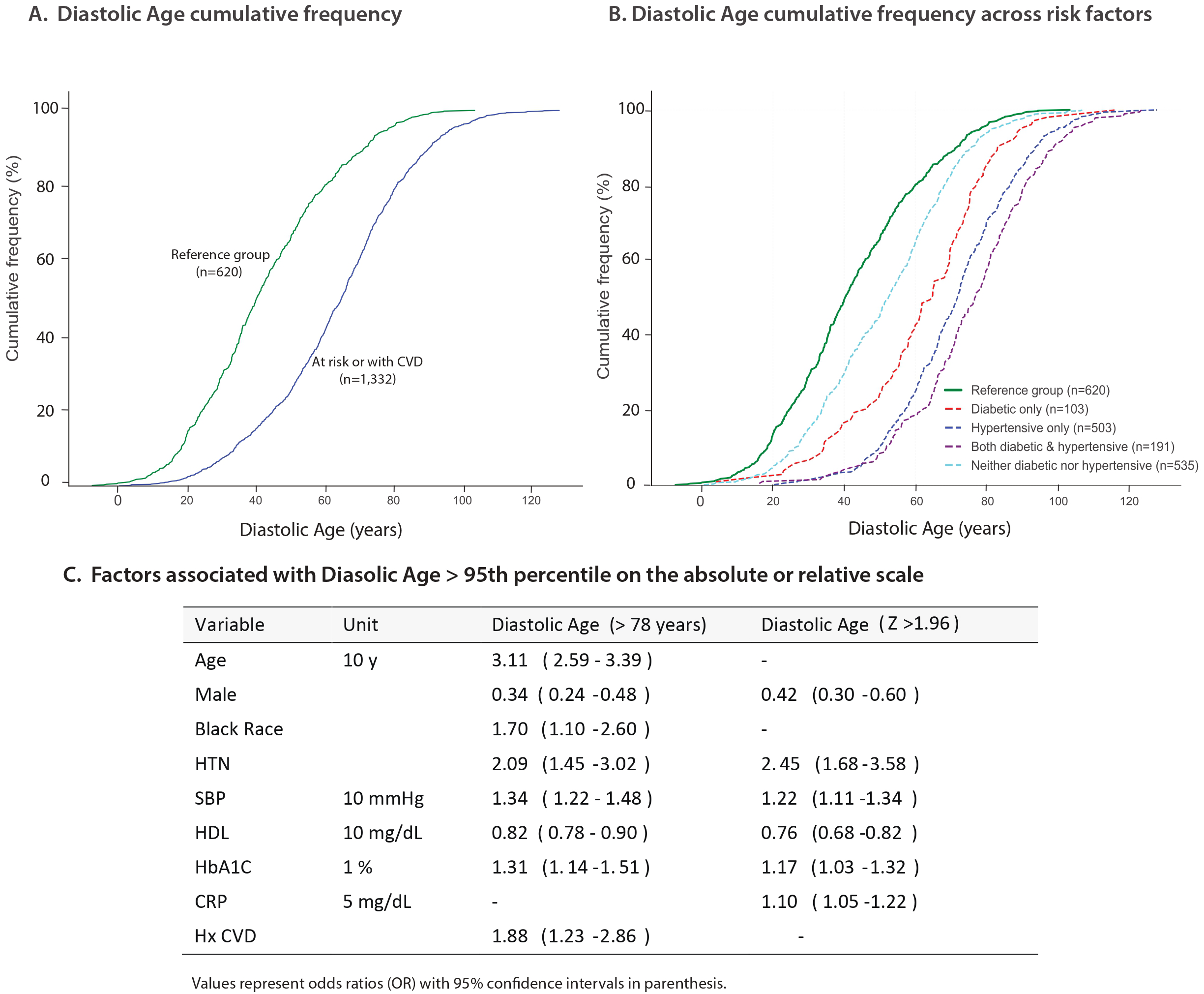
Diastolic Age Distribution and Associated Risk Factors. Cumulative frequency distribution of Diastolic Age comparing the reference group versus higher-risk group (Panel A), cumulative frequency distributions stratified by individual risk factors (Panel B), and odds ratios for clinical factors associated with Diastolic Age > 95^th^ percentile, defined by either absolute threshold (>78 years) or relative Z-score (>1.96) (Panel C). *Abbreviations: CRP, C-reactive protein; HbA1c, hemoglobin A1c (or glycated hemoglobin); HDL, high-density lipoprotein; HTN, hypertension; Hx, history; SBP, systolic blood pressure*.

### LV Diastolic Age and Diastolic Dysfunction

LVDD was identified in 271 participants (8%) of the study cohort. Diastolic Age demonstrated robust discrimination for detecting LVDD, with an AUC of 0.89 (95% CI 0.87–0.92) — superior to the Z-score approach (AUC=0.75, 95% CI 0.72–0.78; **Figure 6A**). Diastolic Age increased progressively across LVDD grades (**Figure 6B**), with the indeterminate grade showing on average higher values than normal.

**Figure 6.**
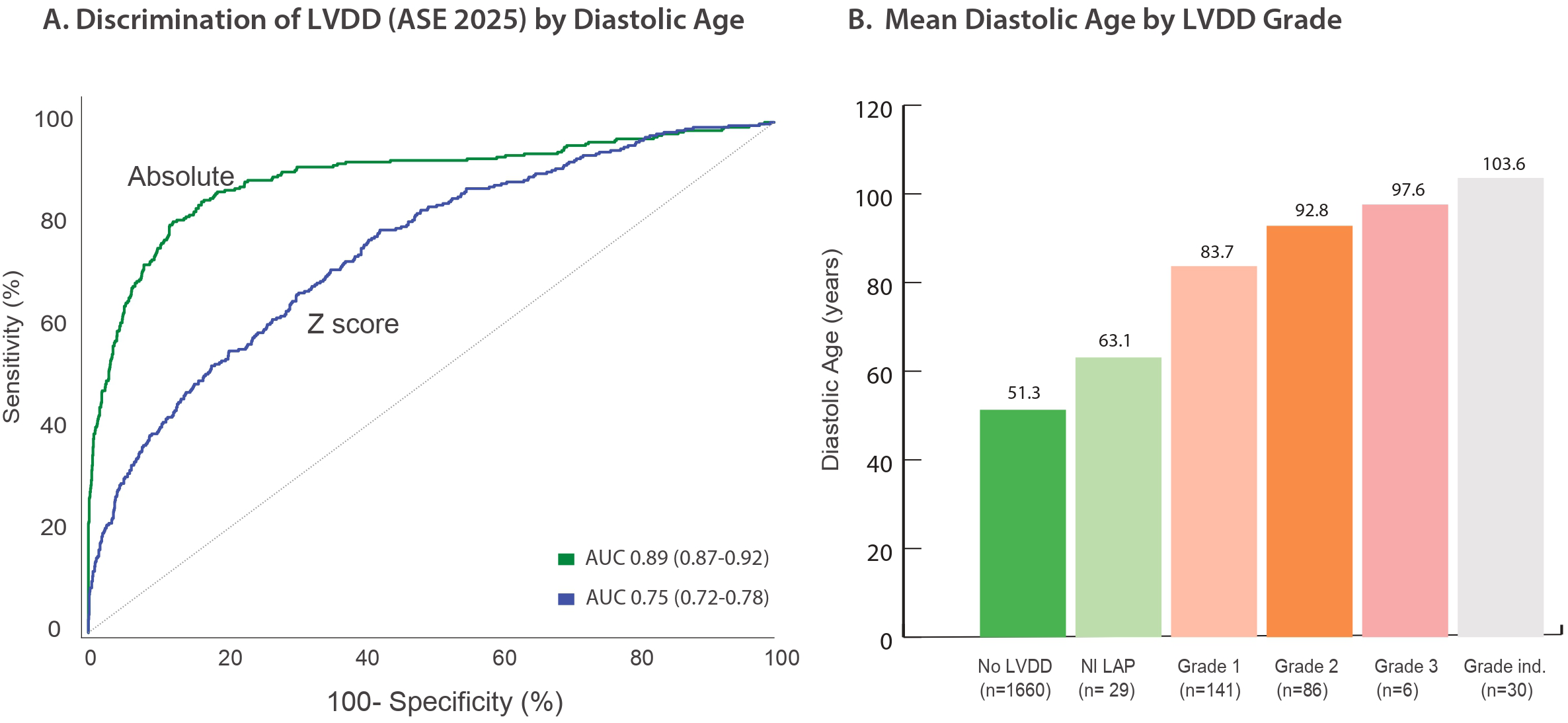
Association of Diastolic Age with Left Ventricular Diastolic Dysfunction (LVDD). Discrimination of ASE diastolic dysfunction classification by Diastolic Age shown by receiver operating characteristic curve (Panel A), and distribution of Diastolic Age across grades of ASE diastolic dysfunction (Panel B). *Abbreviations: Grade ind., Grade indeterminate; Nl LAP, Normal left atrial pressure*.

### Relationship with Epigenetic Clocks

In 1,283 participants with available methylation data, the Levine clock correlated strongly with Diastolic Age (r=0.76, p<0.001; **Figure 7**), whereas the Horvath clock showed a more moderate association (r=0.41, p<0.001; **Supplemental Figure 4**). Residual analyses for both the Levine (r=0.13, p<0.001) and Horvath clocks (r=0.02, p=0.461) revealed substantial scatter around zero, indicating that Diastolic Age captures biological aging dimensions distinct from methylation-based clocks.

**Figure 7.**
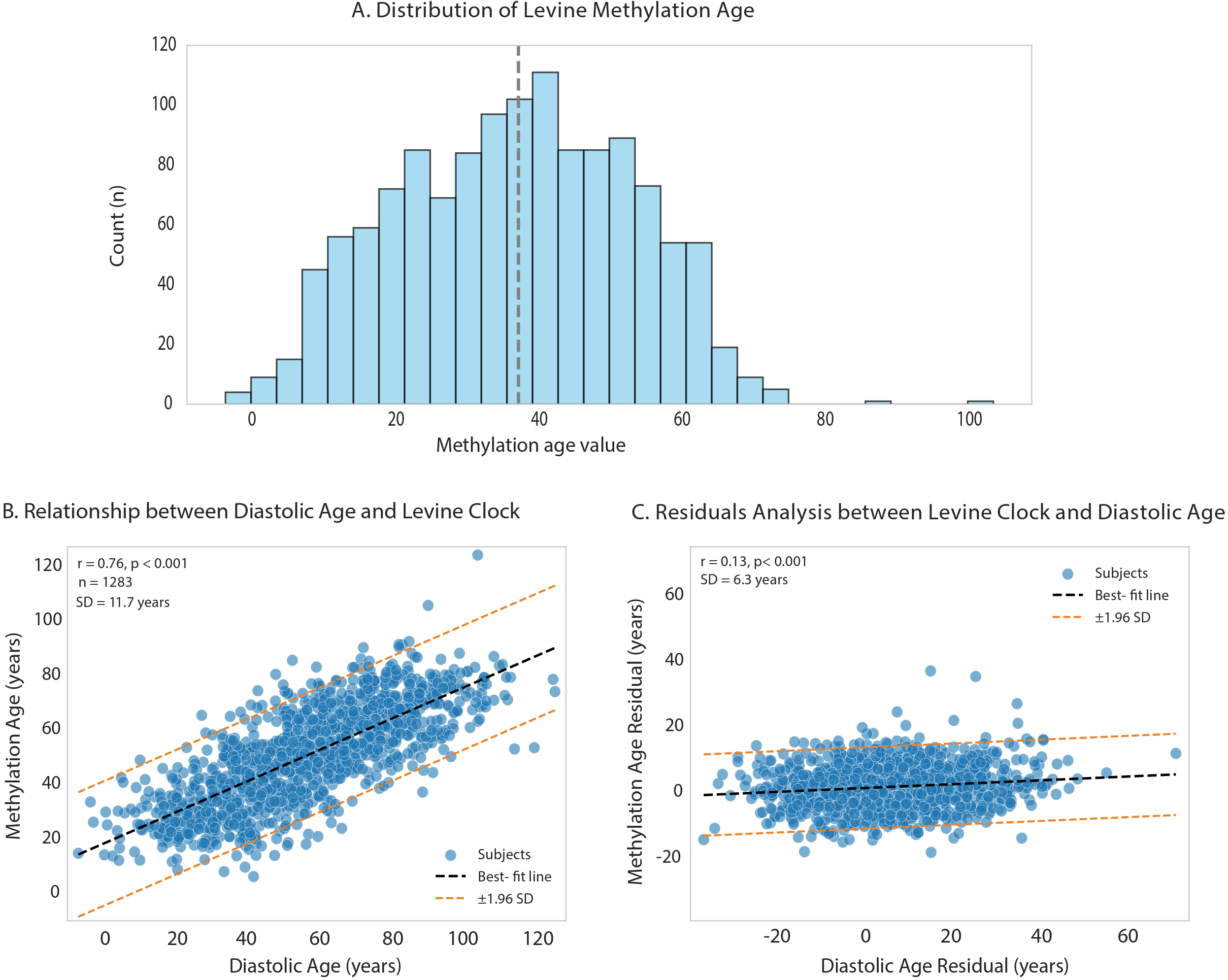
Comparison of Diastolic Age with Levine Methylation Age. Distribution of Levine Age in our cohort (Panel A), correlation with Diastolic Age (Panel B) and residual analysis of the relationship (Panel C).

### LV Diastolic Age and MACE

During a median follow-up of 4.25 years, 98 participants experienced at least one MACE. Of these, 57 events (58%) were atherosclerosis-related, 23 (23%) were new-onset atrial fibrillation, and the remainder comprised thrombotic events (7%), major cardiovascular interventions (5%), incident heart failure (4%), and cardiovascular mortality (2%). Kaplan–Meier curves demonstrated progressively lower event-free survival across higher Diastolic Age quantiles (**Figure 8A**). Survival tree analysis identified Diastolic Age as the principal predictor, with chronological age and sex emerging at shallower tree depths (**Figure 8B**).

**Figure 8.**
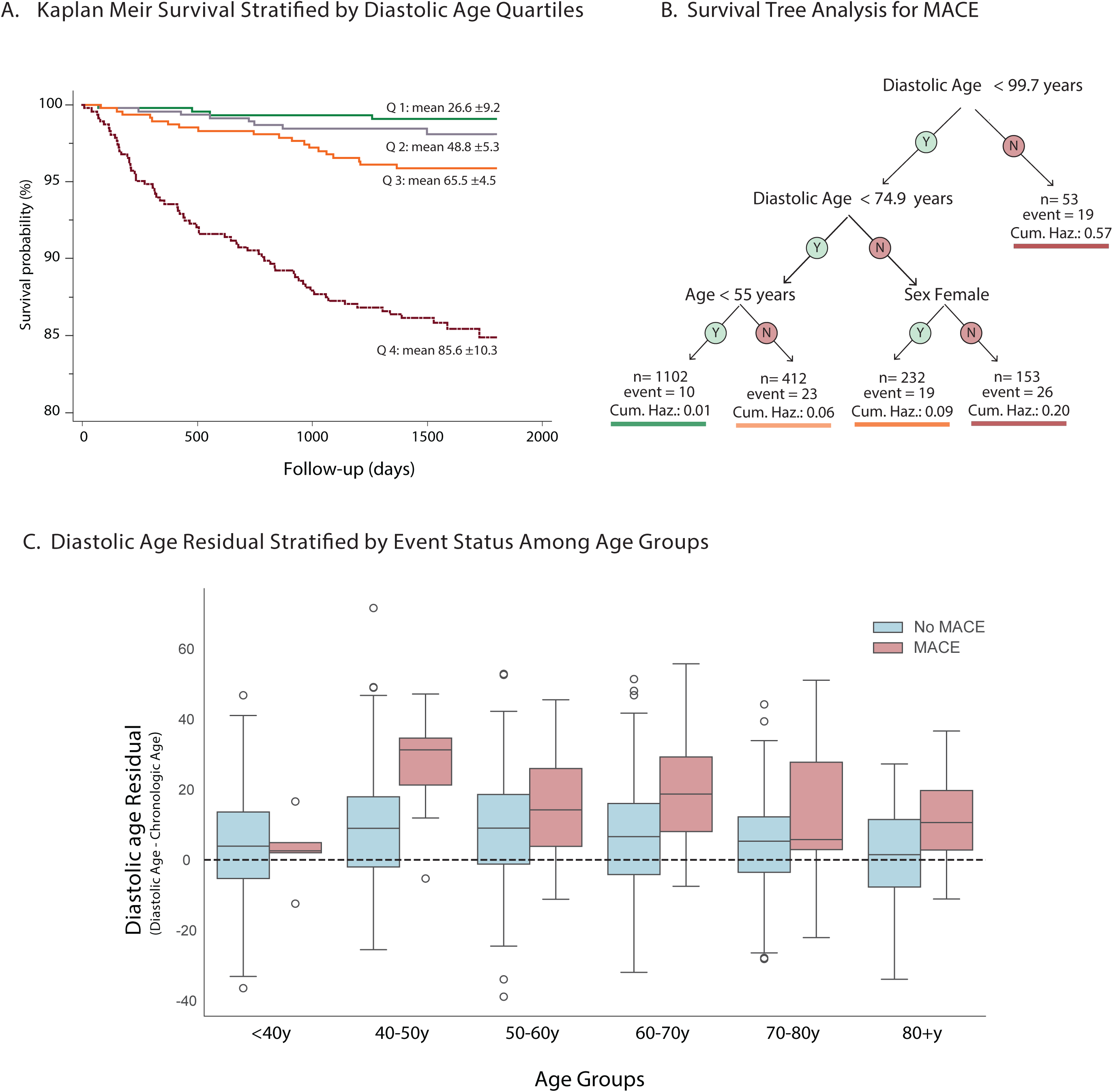
Association of Diastolic Age with Major Adverse Cardiovascular Events (MACE). Kaplan-Meier survival curves stratified by Diastolic Age quartiles (Panel A), survival tree analysis (Panel B), and Diastolic Age residual distribution stratified by event status among age groups (Panel C). *Abbreviations: Cum. Haz.; cumulative hazard*.

In multivariable Cox regression, Diastolic Age remained independently associated with MACE after adjustment for traditional cardiovascular risk factors and preexisting cardiovascular disease (**Table 4**). In a separate model with Diastolic Age and the PREVENT-HF score as sole covariates, both remained independently associated with MACE: Diastolic Age (HR 1.02, 95% CI 1.01–1.04, p=0.008) and PREVENT-HF logit (HR 1.67, 95% CI 1.27–2.19, p<0.001). Participants with MACE consistently exhibited accelerated diastolic aging across all chronological age groups (**Figure 8C**).

**Table 4.** Multivariable Cox Model Predicting Major Adverse Cardiovascular Events (MACE). *SD; standard deviation, NS; non-significant*.

| Variables | Hazard ratio<br>(95% CI) | P-value | -log<sub>2</sub>p |
| --- | --- | --- | --- |
| Diastolic Age (per SD) | 2.30 (1.70-3.19) | <0.001 | 21.62 |
| Age (per SD) | 1.53 (1.09-2.16) | 0.015 | 6.06 |
| Current smoking | 2.63 (1.60-4.30) | < 0.001 | 12.99 |
| History of cardiovascular disease | 2.31 (1.28-3.61) | < 0.001 | 12.08 |
| Male sex | 1.80 (1.19-2.71) | 0.005 | 7.59 |
| Diabetes mellitus II | 1.45 (0.92-2.29) | NS | 3.25 |
| Dyslipidemia | 1.08 (0.69-1.70) | NS | 0.44 |
| Hypertension | 1.07 (0.66-1.72) | NS | 0.33 |

When comparing the prognostic performance of high Diastolic Age (>95^th^ percentile, >78 years) versus LVDD classification by ASE 2025, both models demonstrated excellent discrimination, with C-indices of 0.83 for high Diastolic Age and 0.82 for LVDD diagnosis, and no significant difference between models (ΔC-index = 0.010, 95% CI: -0.015 to 0.031).

### Alternative Outcome-Based Diastolic Clock

To further explore prognostic modeling, we derived an outcome-based diastolic clock by modeling the conditional hazards of age, sex and diastolic parameters. After calibration in the reference group, the resulting equation was: **Outcome-based Diastolic Age = 83.0 – 2.86 (septal e′) – 2.85 (lateral e′) + 1.97 (E/e′ avg) + 0.52 (LAVI) + 3.4 (male).** This outcome-based clock correlated very strongly with the primary PREVENT-HF–based Diastolic Age (r=0.99; **Supplemental Figure 5**).

## Discussion

In this study, we developed and validated “Diastolic Age”; a continuous biological clock that translates routine echocardiographic dara into a quantitative measure of cardiovascular aging. Much like CAC is reported as both an absolute score and an age-based percentile, our model expresses cardiac aging in both absolute years and relative Z-scores. By anchoring the feature weights to the PREVENT-HF risk score, we grounded the metric in established clinical risk. Diastolic Age reliably discriminated ASE diastolic dysfunction and captured a dimension of biological aging that complements standard epigenetic clocks. Furthermore, the exploratory finding that the equation independently predicted MACE highlights its potential prognostic value.

Efforts to quantify biological aging have evolved from molecular markers, such as telomere length and epigenetic clocks, toward accessible, organ-specific models.^2–4,6,8,9,26^ Vascular and cardiac ages derived from CAC^10^, and electrocardiography or echocardiography represent a practical extension of this approach.^11–13,27,28^ Among cardiac measures, LV diastolic function is a particularly compelling candidate.^29,30^ As demonstrated by our network analysis, LV diastolic parameters are strongly associated with chronological age and the PREVENT-HF risk score, making them reproducible and clinically relevant targets for a cardiac-specific aging metric.

A primary challenge in developing biological clocks is selecting an appropriate calibration anchor (**Table 1**). Rather than anchoring to chronological age alone, we weighted echocardiographic features against the validated, sex-specific PREVENT-HF risk score^18^, subsequently calibrating the resulting predictions to chronological age within a healthy reference subgroup. This strategy parallels the development of phenotypic clocks but is specifically optimized for cardiovascular risk.^3^ The stability of this approach was supported by external validation in the WASE and Stanford cohorts, where sex- and race-specific analyses showed consistent performance. Notably, an alternative model calibrated exclusively on incident clinical outcomes yielded near-identical results, suggesting a robust convergence between risk-based and event-based frameworks in evaluating diastolic physiology.

Physiologically, Diastolic Age aligns with expected patterns of biological aging: it is elevated in individuals with traditional cardiovascular risk factors (e.g. hypertension, diabetes) and systemic inflammation (high-sensitivity C-reactive protein), and it increases stepwise with worsening ASE-defined LV diastolic dysfunction.^14,31^ Although Diastolic Age correlated with epigenetic methylation clocks, residual analyses indicated that they capture largely independent aging domains. We hypothesize that epigenetic clocks reflect upstream cellular programming, whereas Diastolic Age captures the downstream functional consequences of cumulative cardiac injury, including mitochondrial dysfunction, altered calcium handling, and fibrotic remodeling.^30,32,33^

From a clinical perspective, the utility of a biological clock depends on its prognostic value and practical applicability. In our exploratory analysis, elevated Diastolic Age was independently associated with MACE, suggesting it reflects a broader cardiovascular risk phenotype beyond isolated heart failure vulnerability. Importantly, high Diastolic Age demonstrated prognostic discrimination non-inferior to established ASE guidelines. By expressing Diastolic Age in dual formats—absolute years and standardized Z-scores—this metric offers a continuous variable to identify accelerated aging, particularly in individuals with borderline or “indeterminate” categorical grades. For patient communication, it translates complex Doppler indices into an accessible metric, potentially facilitating the longitudinal tracking of lifestyle or pharmacological interventions without requiring specialized laboratory assays.

These findings must be interpreted within the context of several limitations. First, while externally validated in the WASE and internal Stanford cohorts, the sample size for our outcome analysis was modest, necessitating evaluation in larger, prospective cohorts. Second, biological clocks are sensitive to measurement variations; inter-center differences in echocardiographic acquisition methods (e.g., Doppler peak versus modal frequency) could influence parameter estimates. Third, excluding patients with atrial fibrillation or pacemakers likely underestimates the true burden of diastolic aging, as these arrhythmias are often manifestations of advanced diastolic dysfunction. Future studies should establish boundary constraints for extreme age values and explore multimodal approaches, integrating electrocardiographic data and biomarkers (e.g., NT-proBNP) to further refine age estimation.

study has several limitations. Although the model was validated in two independent cohorts, the overall sample size, particularly for outcome analysis, remains modest. As with all biological clocks, calibration across diverse populations and imaging settings remains a key challenge. This is especially relevant for echocardiography, where inter-center differences in acquisition methods (e.g., Doppler peak versus modal frequency) may affect parameter estimates. The exclusion of patients with atrial fibrillation or pacemakers may underestimate the burden of diastolic aging in real-world populations and introduce selection bias, as arrhythmias and conduction system disease can themselves be manifestations of advanced LVDD. Future studies should evaluate the model’s generalizability in older and/or sicker patient populations. Additional constraints may also be needed to bound extreme or negative Diastolic Age values. Future studies could also integrate deep learning approaches and ensemble models combining echocardiography, electrocardiography, and biomarkers such as NT-proBNP to further refine and individualize Diastolic Age estimation.

In conclusion, Diastolic Age translates routine echocardiographic measurements into a continuous, quantitative marker of cardiac aging. By complementing categorical diastolic dysfunction grading and molecular epigenetic clocks, it may offer a practical, non-invasive tool for cardiovascular risk stratification and longitudinal patient assessment.

## Data Availability

Data is available upon reasonable requests.

## Disclosures

KMA has received consulting fees from Alexion, Alnylam, Bayer, BridgeBio, Novo Nordisk, and Pfizer. SAS report employment and equity ownership in Verily Life Sciences. SSSO holds is a consultant and board member of CytoReason and holds equity. AK serves as Chief Medical Officer of Argus Cognitive, Inc., unrelated to the present manuscript. MMD is Director of the DCRI Multimodality Core Laboratory, which received institutional support from Verily for the conduct of the PBHS. FA is Director of an academic core lab with institutional (MedStar Health) research contracts/grants/collaborations with <u>Us2.ai</u>, Ultromics, egnite, Tomtec, MyCardium, Abbott, Edwards, BSC, Medtronic, Ancora Heart, Neovasc, Innovheart, Polares Medical, Foldax, Intershunt, CorFlow, J&J, Aria CV, Corcym, Xeltis, Tricares, Vdyne, Croivalve, Sanofi, Novartis, BMS. KWM has received research grants from Google/Verily. All other authors do not report any disclosures or competing interests.

## Funding

The Project Baseline Health Study was funded by Verily Life Sciences, Dallas, Texas. Analysis for this projects was funded by Stanford Cardiovascular Institute.

## Registration

The Project Baseline Health Study is registered at https://ClinicalTrials.gov (Identifier: NCT03154346).

## Acknowledgements

In memory of our dear steadfast mentor and friend Dr. Roberto M. Lang who passed away unexpectedly.

## Notes

### Competing Interest Statement

The authors have declared no competing interest.

### Funding Statement

The Project Baseline Health Study was funded by Verily Life Sciences, Dallas, Texas. Analysis for this project was funded by Stanford Cardiovascular Institute.

### Author Declarations

The study was approved by the Stanford University and Duke University Institutional Review Boards

